# Emergent Heterogeneity in Compartmental Disease Models

**DOI:** 10.64898/2026.11.13.26367950

**Authors:** Michael Roswell, Azadeh Aghaeeyan, Tapan Goel, Stephen J. Beckett, Joshua S. Weitz, Jonathan Dushoff

## Abstract

Heterogeneity in infectious disease spread reshapes the risk and severity of outbreaks. This heterogeneity can arise from intrinsic individual-level differences in susceptibility, contact patterns, and infectious durations. Heterogeneity can also emerge from disease dynamics in a canonically homogeneous population, due to the fact that individuals randomly become infected at different times, recover after different intervals, and have access to different numbers of susceptible individuals. Here, we explore the mechanisms generating emergent heterogeneity by analyzing secondary case distributions within the SIR model. We show numerically and prove analytically that the variation in secondary cases, calculated over the course of the epidemic, does not change with *R*_0_. This is in contrast to the increasing variation in secondary cases in the linearized SIR model, often used to study heterogeneity at the beginning of an outbreak. When we consider “forward cohorts” of individuals infected at the same time, between-cohort variation increases as *R*_0_ increases, while within-cohort variation decreases. We show that these effects balance exactly, leading to the invariance of the variation in secondary cases with respect to *R*_0_. These results demonstrate the importance of considering emergent, as well as intrinsic, variation between individuals and of considering heterogeneity of transmission spread over the whole epidemic cycle, including substantive dynamical effects that appear even in the initial outbreak phase.

## 1 Introduction

Real outbreaks have significant heterogeneity in the number of secondary cases – including superspreading, marked by a small fraction of individuals causing a disproportionately large fraction of realized transmission [12, 19]. Super-spreading can help explain why epidemics take off in some situations and not in others, despite apparently similar contexts [4, 11]. Leveraging knowledge of superspreading can make control efforts more effective [5, 14, 18], especially at the beginning of outbreaks [6]. Correspondingly, theoretical treatment of super-spreading has emphasized individual-level transmission patterns while assuming a fully susceptible population [9, 12]. Models of superspreading often focus on linking observed secondary case counts to a continuous random variable, the *individual reproductive number*, via a Poisson sampling process. Canonically, the individual reproductive number is modeled as a gamma distribution, giving rise to negative binomial secondary case distributions [12]. In this way, variation arising from the stochastic transmission process is conceptually separated from *intrinsic heterogeneity* in expected infectiousness that may emerge from potential differences in biology or behaviors between individuals [1, 3, 7].

Conventional models accounting for heterogeneity typically assume a constant pool of susceptible individuals, implicitly or explicitly focusing on the very beginning of an outbreak. Over longer timescales, susceptible depletion and replenishment drive population-scale disease dynamics. Changes in the size of the susceptible population change the proportion of contacts that lead to successful disease transmission and drive differences in expected cases between different cohorts of infectors. Many deterministic epidemiological models also include *explicit heterogeneity*, for example by adding compartments for different demographic or geographic groups [17], which can have different average rates of mixing, transmission, or recovery. When implemented in terms of individual events, the inclusion of explicit heterogeneity reproduces superspreading in linear compartmental models [13]. Despite shared theory motivating models that highlight individual-level transmission, and those that emphasize population-level transmission dynamics, a gap persists in how each model framework treats individual-level transmission heterogeneity.

The canonical SIR (Susceptible-Infected-Removed) model uses ordinary differential equations (ODEs) to describe disease dynamics. The SIR model assumes constant, uniform infectivity and recovery times across all individuals during the infectious stage [8, 10]. Although it appears homogeneous, even the canonical SIR model has two sources of *emergent heterogeneity*. First, the changing effective reproductive number arising from susceptible dynamics drives variation between fractions of the population infected at different times. Second, the exponentially distributed recovery times assumed implicity by the ODE framework drive variation in the number of infections attributable to fractions of the population infected at a given time. In other words, the deterministic SIR model implies the presence of variation in the expected number of secondary cases.

Here we delineate sources of individual variation in transmission inscribed in simple dynamical models. We trace how these sources compose into observed or observable heterogeneity in different settings, for example contrasting how heterogeneity emerges in dynamical models versus linearized, or branching-process models. We begin by revisiting early findings that emergent heterogeneity can drive variation in the secondary case distribution, and discuss how individual-level heterogeneity has a stronger effect when the basic reproductive number *R*_0_ is smaller. We then show, first via simulations and then analytically, that emergent heterogeneity in secondary cases is not affected by *R*_0_ across a full SIR outbreak. Throughout, we highlight the understudied, but critically important, role of disease dynamics *per se* in generating individual-level heterogeneity. We aim to spur the development of theory and methods that better integrate heterogeneity and dynamics, ultimately facilitating feedback between data on incidence and individual-level transmission chains, and mechanistic model specification, identification, validation, and intervention design.

## 2 Results

### 2.1 Heterogeneity in the secondary case distribution in the linearized SIR model

The early outbreak phase of an epidemic is modeled using a linearized SIR model, which assumes that the pool of susceptible individuals does not change appreciably. Early dynamics can thus be characterized by a transmission rate and an infectious period. Demographic stochasticity generates heterogeneity in this model despite the lack of explicit accounting of individual-level variation in transmission rates and infectious periods. Since recovery is Markovian, the model implicitly assumes an exponentially distributed infectious period, resulting in an exponentially distributed expected infectiousness (or *individual reproductive number*) with mean *µ* = *R*_0_ and generalized dispersion *κ* = 1. The resulting distribution of secondary cases is geometric and has a mean equal to *R*_0_ (see Box 1). Thus, the expected and realized secondary-case distributions are characterized by *R*_0_ (Figure 1a).

**Figure 1:**
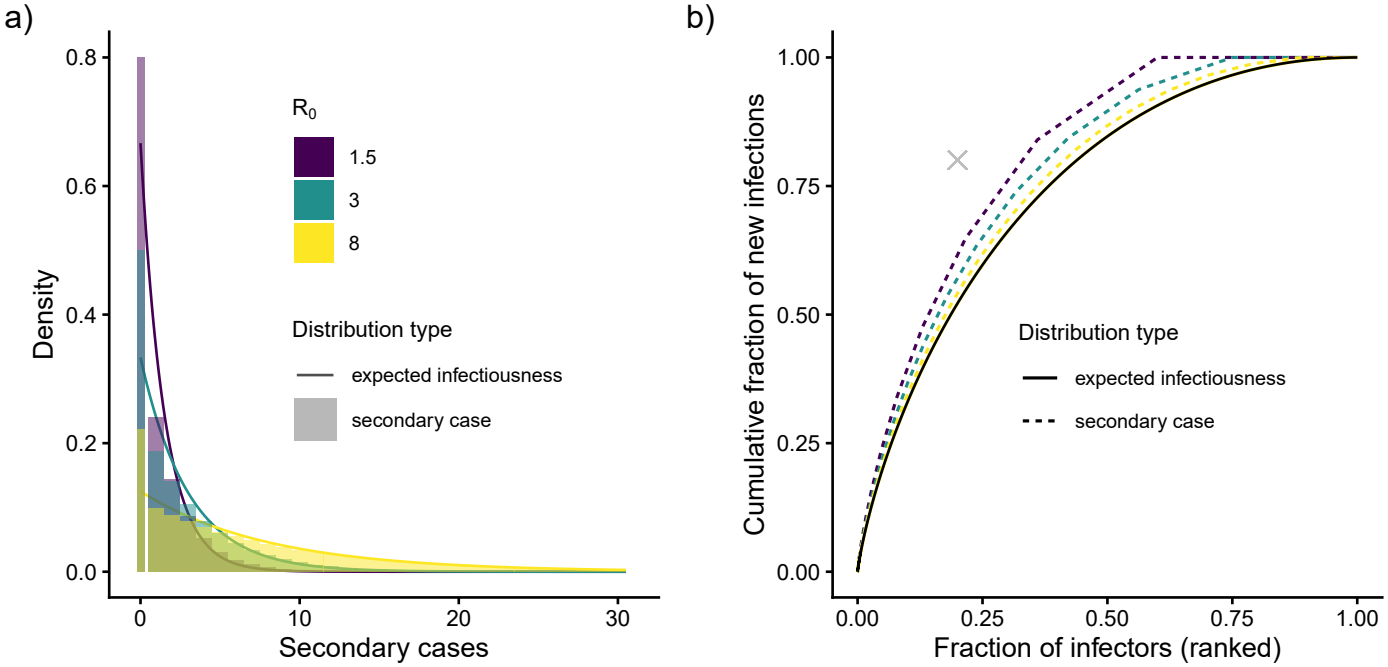
Heterogeneity in the linearized SIR model due to implicit variation in recovery times among infectors. (a) Both expected infectiousness (density curves) and secondary case distributions (density histograms) for the outset of an SIR epidemic, where the population is fully susceptible. (b) Inequality curves, defined as the cumulative fraction of new infections versus the cumulative fraction of infectors. Curves (in black) for the expected infectiousness across varying *R*_0_ coincide. Inequality curves derived from the secondary case distributions are shown with dashed lines. The cross marks “20-80” point, where 20% of infectors cause 80% of infections. In panel a, because the first bin (at zero) sits at the boundary of support for each distribution, we have plotted this bin as double the density and half the width; this adjustment preserves area-to-area correspondence with the probability density function, while facilitating visual comparison of the heights of the density and mass functions.

#### Box 1. Linking variance across discrete and continuous time models

During the early stages of an outbreak, realized secondary-cases counts are typically modeled using a negative-binomial distribution, with mean *µ* and shape, or “dispersion,” parameter *k*. This distribution corresponds mechanistically to a gamma-distributed “expected number of (secondary) cases”, with the same mean and shape parameters. The “realized number of (secondary) cases” is then given by the convolution of this expected number with a Poisson distribution (reflecting necessary, individual-level stochasticity arising from a Poisson contact process) [12].

This is illustrated in Figure 1 for the case where differences in infectiousness arise from random recovery times under simple SIR assumptions, and is thus exponentially distributed. Expected infectiousness scales exactly with *R*_0_, so that the smooth curves in Figure 1b, are all the same, but the relative variance in realized cases increases when *R*_0_ is small, because the proportional amount of randomness added by the Poisson step is larger.

We note a couple of points. First, the squared coefficient of variation (SCV) of the underlying Gamma distribution is *κ* = 1*/k*, so its variance is 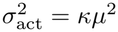 . The resulting variance in secondary-case counts is *σ*^2^ = *κµ*^2^+*µ*, with the last term arising from the Poisson step of the negative-binomial construction. In the case of Figure 1, exponentially distributed infectiousness (corresponding to a geometric distribution of secondary cases), *κ* = *k* = 1. Second, the relationship between 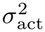 and *σ*^2^ holds regardless of the form of the expected infectiousness distribution, due to the Law of Total Variation [16].

We therefore suggest *κ* as a generalized dispersion parameter. This has the advantages of (a) less dependence on a specific assumption about distribution (and parameterization), and (b), the intuitive idea that variation increases when “dispersion” increases.

The heterogeneity in the secondary case distribution means that a large fraction of the initial secondary cases are generated by only a few infectors, even when starting from the canonical homogeneous assumption. The proportion of *expected* cases caused by the individuals who remain infectious the longest is the same for any value of *R*_0_, but the proportion or *realized* cases caused by the most effective infectors is larger for smaller values of *R*_0_, since the relative importance of the Poisson infection process increases (Figure 1b). As *R*_0_ becomes smaller, an increasing fraction of infected individuals generate no secondary infections, so the individuals who do transmit make up a smaller fraction of all infectors, such that realized heterogeneity is expected to increase as *R*_0_ decreases.

### 2.2 Heterogeneity in secondary cases over an epidemic appears independent of *R*_0_

Next we consider the impact of the depletion of the susceptible population during outbreaks on secondary case distributions. We simulated stochastic SIR epidemics with different *R*_0_ values in a population of 10^4^ individuals. For each infected individual, we recorded the number of secondary cases they generated during the outbreak. Figure 2 shows realized distributions of secondary cases caused by individual infectors over the entire outbreaks. Unlike in the linearized scenario shown in Figure 1, where the susceptible pool remains fully populated, the secondary case distributions, computed across the entire outbreak, appear indistinguishable across a broad range of the key parameter *R*_0_ (compare panel (a) of Figure 1 with the entire set of panels in Figure 2).

**Figure 2:**
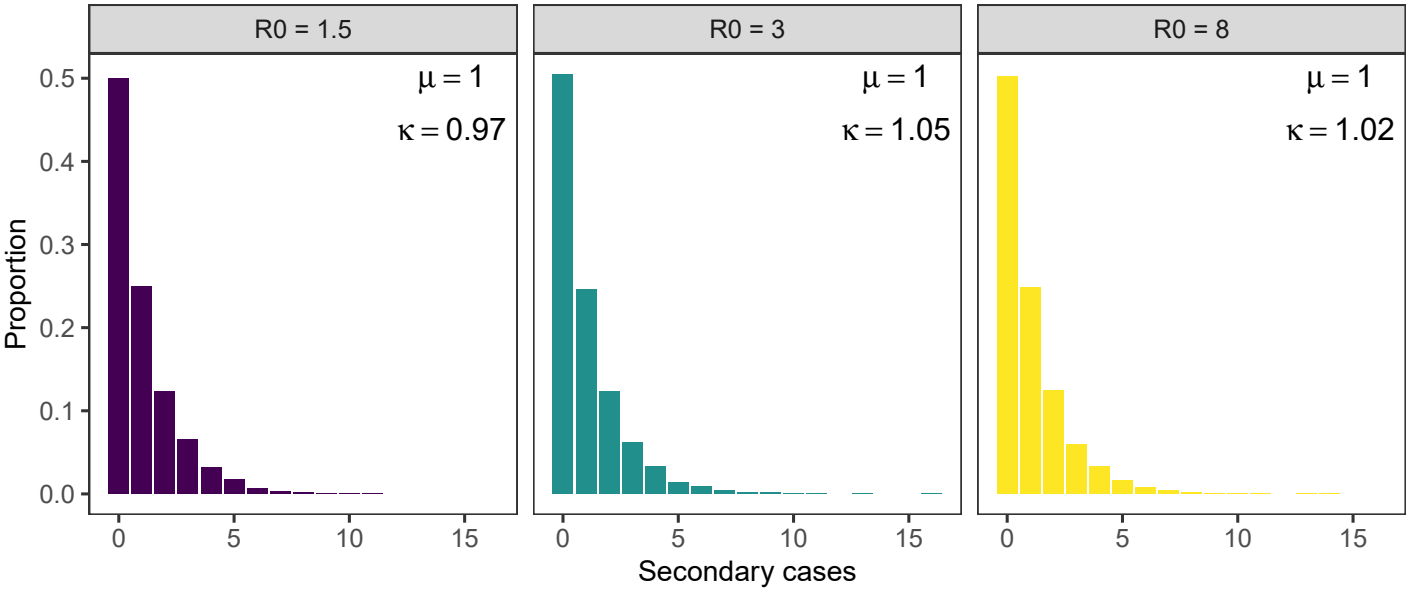
Heterogeneity in the number of secondary cases over an entire epidemic appears to be independent of *R*_0_. Realized secondary cases collected from individual-based simulations of the SIR model for *R*_0_ = 1.5, 3, and 8 (from left to right respectively); *µ* and *κ* denote the mean and generalized dispersion, respectively, of the realized case distribution obtained from a single realization, conditional on a large outbreak occurring, of the individual-based model for each *R*_0_, in a population of 10^4^ individuals with one initial infectious individual.

Two key features of the secondary case distributions across an entire out-break are salient. First, the mean of the secondary case distribution is approximately 1, as expected. Since we are considering a single outbreak seeded by a single initial case, the number of infectors must almost equal the number of infectees. Second, the variance of the realized secondary case distribution is approximately 2, corresponding to a variance of 1 in the expected case distribution, independent of *R*_0_. This result is unexpected. In the absence of susceptible depletion, heterogeneity in expected secondary cases is constant across values of *R*_0_ (Figure 1b). During the full epidemic, we expect large-*R*_0_ simulations to cause large dynamical changes, leading to larger variation in expected (and thus realized) secondary cases. At first glance, it is not clear what could balance the dynamical variation in susceptibles for different values of *R*_0_ to leave the variance constant.

### 2.3 Heterogeneity in secondary cases over an epidemic is independent of *R*_0_, a proof

We set out to explore the basis for the observation that both the mean and variance of realized cases in the stochastic SIR model appear to be invariant with respect to changes in *R*_0_. To do so, consider the nondimensionalized, deterministic ODE-based SIR model:

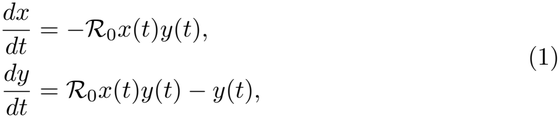

where *x*(*t*) is the proportion of susceptible individuals at time *t, y*(*t*) is the proportion of infectious individuals at time *t, t* is time measured in units of the mean infectious duration and *R*_0_ is the basic reproduction number.

To calculate the mean and variance of the secondary cases, we must track infections and the infectors that caused those infections. While individual infectors and cases cannot be tracked in this deterministic SIR model, we can track cohorts of infectors and the cases caused by them. By calculating the weighted moments of the expected cases caused by each cohort, we can obtain the heterogeneity of the secondary cases over the epidemic.

We define an “interval cohort”, 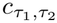, as the fraction of infectors that become infectious at time *τ*_1_ and stop being infectious at time *τ*_2_. The weight of the cohort, *w*(*τ*_1_, *τ*_2_), is the number of infectors in the cohort. The number of infectors who become infectious at time *τ*_1_ is *i*(*τ*_1_) = *R*_0_*x*(*τ*_1_)*y*(*τ*_1_). This quantity, *i*(*τ*_1_) is also called the incidence of infection, or the number of new cases that appear at time *τ*_1_. Only a fraction of these infectors remain infectious till time *τ*_2_. If *f* (*·*) denotes the probability density function of the infectious period, then

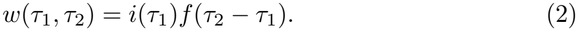

Now that we have the cohort weights, we need to calculate the number of secondary cases caused by an interval cohort. The total number of cases caused by the interval cohort 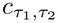 over the course of the epidemic is

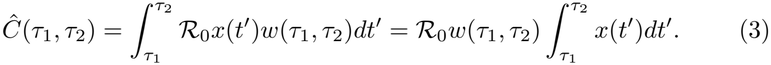

Then the expected number of secondary cases caused by individuals in cohort 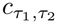 is

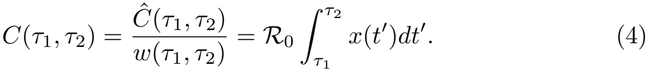

Given the number of expected secondary cases and weight for each cohort, we can calculate the moments of the distribution of expected secondary cases. The *k*^th^ weighted raw moment of this distribution is

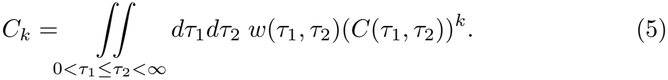

With this definition of the raw moments, the assertion that the heterogeneity of the secondary case distribution over the entire epidemic is invariant is equivalent to:

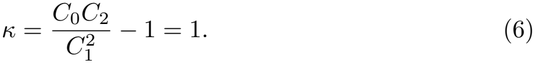

We calculate the moments and confirm that the above assertion is true in section S.1.1. We note that the result follows from two assumptions: the distribution of the infectious duration is memoryless, and the initial infectious cohort is infinitesimal, i.e., *y*(*t* = 0) *→* 0. The first assumption is true for the canonical SIR model where the duration of infectious periods is exponentially distributed. We discuss relaxing the second assumption in section S.1.2.

### 2.4 Offsetting contributions of within- and between-cohort variance drive *R*_0_-invariance of heterogeneity in secondary cases

To understand why the heterogeneity in secondary cases over the course of the epidemic is independent of *R*_0_, we divide the population into “forward cohorts” of individuals infected at the same time, and use variance partitioning to split the heterogeneity into two components: a within-cohort component and a between-cohort component. Individuals within the same forward cohort have access to the same susceptible pool initially but have differences in their secondary case counts as a result of their exponentially distributed infectious periods. Individuals in different forward cohorts also differ in the starting size of the susceptible pools that they have access to. It is important to note that both early and late cohorts have the same distributions of infectious periods and expected infectiousness but due to outbreak dynamics ((Figure 3a) that deplete the susceptible pool over time (Figure 3b), later cohorts, on average, have much lower secondary cases than earlier cohorts. Thus, the between-cohort variance can be attributed to susceptible depletion.

**Figure 3:**
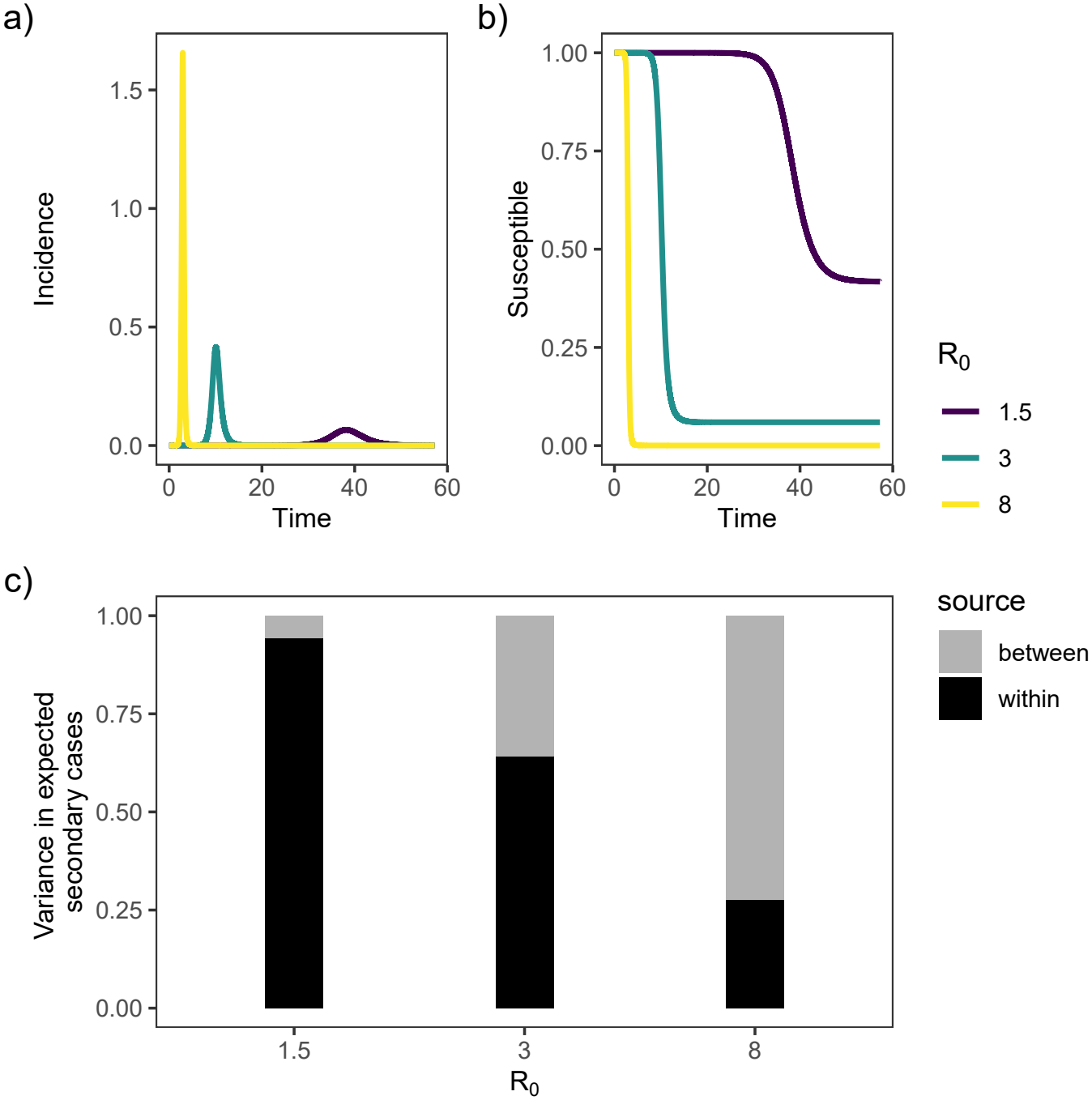
Deterministic SIR dynamics and within- and between-cohort variance. (a) Incidence and (b) proportion of susceptibles as functions of time, for different values of *R*_0_. The epidemics were modeled using an ODE-based SIR model. Time is in units of mean infectious period. (c) Stacked bar plot showing within- and between-cohort components of variance in secondary cases calculated numerically for different values of *R*_0_.

Numerical calculation of the within- and between-cohort components of the variance in secondary cases using the ODE-based SIR model (see methods for details of the calculation) shows that as *R*_0_ increases, the between-cohort component of the variance increases and the within-cohort component of the variances decreases (Figure 3c). However, the sum of the two components, remains constant across *R*_0_. As *R*_0_ increases, the variance in the size of the forward cohorts (Figure 4a) and well as the speed and severity of susceptible depletion also increases (Figure 3b). As a result of the susceptible depletion, the mean number of secondary cases also decreases rapidly across forward cohorts (Figure 4b), with earlier forward cohorts having a much higher mean of secondary cases compared to later forward cohorts. Further, the decline in the mean number of secondary cases across forward cohorts becomes sharper as *R*_0_ increases. As a result, the between-cohort variance in the number of secondary cases increases with *R*_0_.

**Figure 4:**
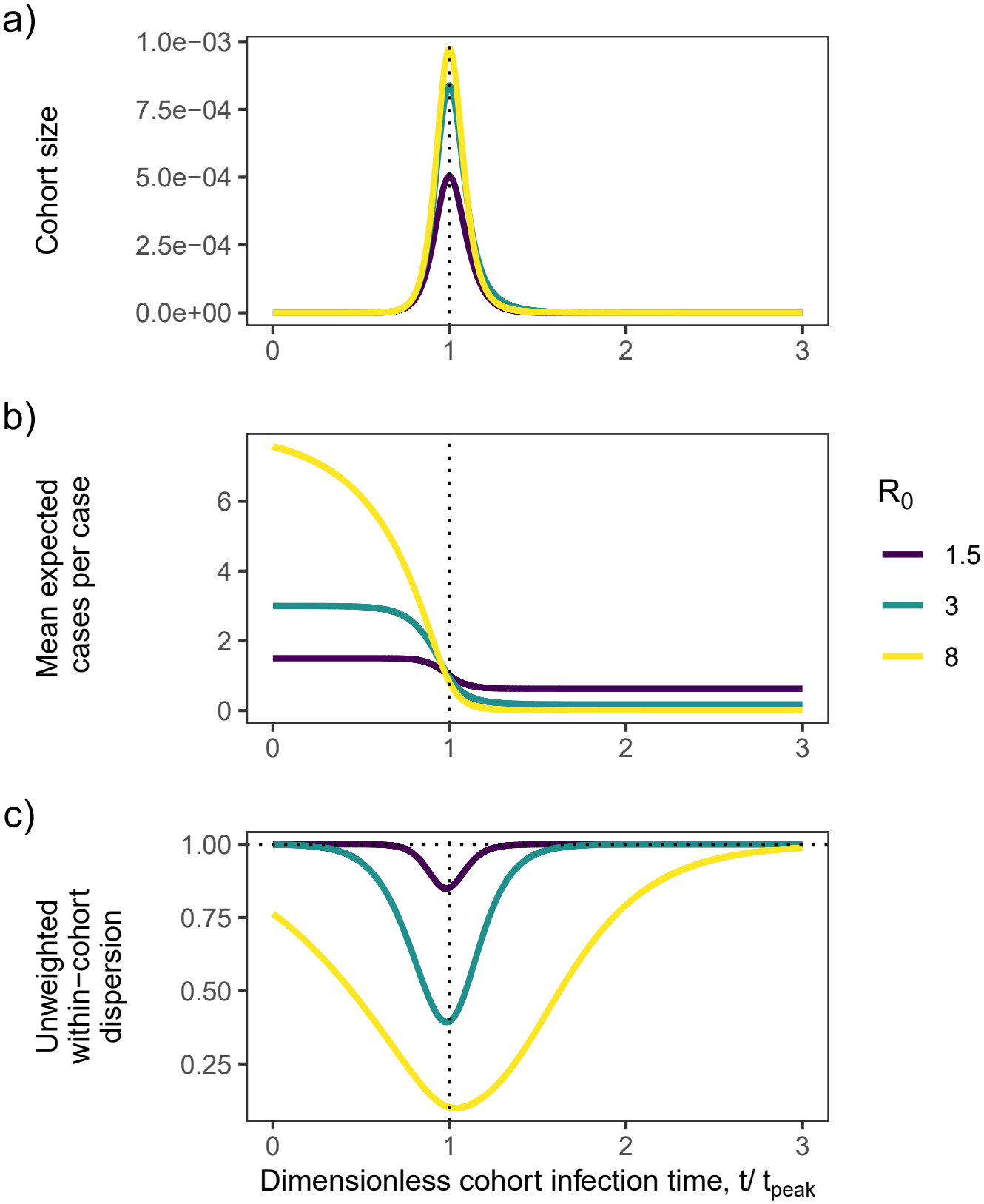
Expected secondary cases of forward cohorts. (a) Relative cohort size as a function of time of infection. Time has been normalized by the time of peak incidence (*t*_peak_). Individuals infected during [*t, t* + Δ*t*] form a cohort, with Δ*t* = 2 *×* 10^*−*4^*t*_peak_. (b) Mean expected secondary cases (cases per case) for each cohort. (c) Within-cohort dispersion of secondary cases. Note that the x-axes refer to dimensionless cohort infection time. For example, the mean expected secondary cases at *t/t*_peak_ = 1 does not refer to the mean expected secondary at time *t* = *t*_peak_. Instead, it refers to the mean expected secondary cases of the cohort that became infectious at time *t* = *t*_peak_.

The within-cohort variance, on the other hand, decreases as *R*_0_ increases (Figure 3c). Since the mean number of secondary cases varies widely between early and late cohorts, we look at the within-cohort dispersion in secondary cases to better understand how within-cohort heterogeneity changes between early vs late cohorts for an epidemic with a given *R*_0_. For a given *R*_0_, early cohorts have high dispersion in secondary cases, driven by access to a large susceptible pool (Figure 4c). As the epidemic progresses, the susceptible pool depletes (Figure 3b) and the within-cohort dispersion in secondary cases decreases (Figure 4c). The within-cohort dispersion increases for the later cohorts, driven by the shrinking within-cohort mean in the number of secondary cases (Figure 4c). However, the within-cohort contribution to the total variance depends not just on the within-cohort dispersion, but also on the cohort weight. Once the contributions of each forward cohort to the variance are appropriately weighted by the cohort mean and cohort size, we find that the weighted within-cohort dispersion is low for both early and late cohorts, and is high for cohorts that arise in the middle of the epidemic (Figure 4a) for a given *R*_0_. Indeed, across different values of *R*_0_, we find that the weighted within-cohort dispersion decreases as *R*_0_ increases (Figure 3c). Together, the decrease in the within-cohort variances balances the increase in the between-cohort variance, leading to the total variances in secondary cases being independent of *R*_0_.

## 3 Discussion

Quantifying heterogeneity in secondary-case distributions helps explain how epidemics unfold and how they can be contained. Although there have been efforts to quantify individual-level heterogeneity in contact rates [19] and to parameterize biologically realistic secondary case distributions, abstracted from the mechanism [12], these efforts have been largely separate from the construction and evaluation of the compartmental models that enable epidemic forecasting and intervention design (but see [13]). Here, we expand the bridge between individual- and population perspectives of heterogeneity by studying epidemiologically-relevant cohorts of infectors that become infectious at the same time.

We first showed how demographic stochasticity and a Poisson contact process result in heterogeneity in secondary cases, even in the linearized SIR model with its canonical assumption of homogeneity between individuals, and lack of susceptible depletion. Next, we showed how heterogeneity emerges in the SIR model of disease dynamics. Depletion of susceptibles over the course of an epidemic leads to differences in realized secondary cases between early and late infectors, despite sharing identical underlying infectiousness distributions. We then found, via simulations, and proved a surprising result – that the total variance in secondary cases over an epidemic is constant across all values of *R*_0_. This invariance is unexpected: in the linearized model, relative variance within cohorts is the same across values of *R*_0_, so we expected the larger between-cohort differences when *R*_0_ is large to drive larger total variance in that case. Instead, we found the susceptible depletion also drives a corresponding decrease in within-cohort variance.

Our cohort-level analyses reveal the importance of susceptible depletion, even in early stages of the epidemic. We find that even very early cohorts of infectors have mean secondary case counts much below the theoretical *R*_0_ (Figure 4b). This discrepancy becomes larger as *R*_0_ increases. In practice, this means that mean secondary case counts systematically underestimate *R*_0_, especially when it is appreciably greater than 1 [15]. We also showed that the dispersion parameter *κ* can unify estimates of heterogeneity, without reliance on a particular distributional assumption, across individual-based and compartmental models, particularly beyond the initial outbreak phase, facilitating comparisons with data measured at the individual level.

The independence of the variance in secondary cases hinges on the SIR model structure. It requires that the infectious period be exponentially distributed. Further, it requires a well-mixed population. However, our results with the SIR model provide a baseline for comparing heterogeneity in secondary cases across different compartmental models. More broadly, by making the implicit explicit, the framework developed here provides a pathway to estimate and incorporate sources of emergent heterogeneity that arise from the dynamics of disease spread, and eventually to combine them with intrinsic individual-level heterogeneity.

## 4 Materials and Methods

### 4.1 Individual-based SIR model

We simulated an individual-based SIR model for a population of *N* individuals. Each individual was assigned a numerical index and an infectious duration, drawn from an exponential distribution with rate *γ*. Next, we built a matrix containing all possible pairs of individuals, and generated a pool of contact events by randomly sampling pairs from this matrix with replacement. The contact rate for each pair of individuals is *β/*(*N −* 1), where *β* is the transmission rate. The total rate of contact events in the population was obtained by multiplying this rate by the total number of possible pairs, *N* (*N −* 1)*/*2, giving *βN/*2. The time interval between successive contact events was then drawn from an exponential distribution with a rate *βN/*2.

Throughout the simulation, we tracked the infection status and number of infections caused by each individual. We initialized the simulation by assigning a single random individual to be infectious. We then ran the simulation forward in time, with successive time steps corresponding to successive contact events. At each time step, we checked the infection statuses of the individuals that came in contact based on when they became infectious and their assigned infectious periods. If both individuals were susceptible or infectious or recovered/removed, we moved to the next time step. If one of the individuals was recovered/removed, we moved to the next time step. However, if one of the individuals was infectious and the other was susceptible, we changed the infection status of the susceptible individual to infectious, noting the time when they became infectious. We also increased the tally of secondary cases caused by the infectious individual by 1. Then, we moved to the next time step. We stopped this process when either no infectious or no susceptible individuals remained.

We used this simulation scheme to generate Figure 2. We set the mean infectious period 1*/γ* to 1 and computed the number of secondary cases in a population of size *N* = 10^4^.

### 4.2 Deterministic SIR model

The deterministic ODE-based SIR model is:

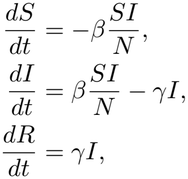

where *S*(*t*), *I*(*t*), and *R*(*t*) are the numbers of susceptible, infectious, and recovered/removed individuals at time *t*, respectively. We denote the transmission rate by *β* and the recovery rate by *γ*. The population size is *N* = *S* + *I* + *R*, which is constant, so we do not need to track *R* explicitly. We then rewrite the equations in terms of the following rescaled variables: the proportion of susceptible individuals, 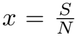 ; the proportion of infectious individuals, 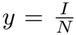 ; and time measured in units of mean infectious period, 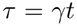 . The rescaled model is:

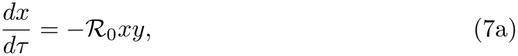

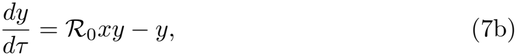

where 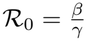 is the basic reproduction number.

### 4.3 Calculation of heterogeneity in the linearized SIR model

To calculate the density histograms for a secondary case distribution, we computed the geometric probability mass function with mean *R*_0_ at points 0, 1, …, 30. We used the convention that the geometric distribution counts the number of failures before the first success (Figure 1a, density histograms). To calculate the density curves for the expected infectiousness, we evaluated the exponential probability density function with mean *R*_0_ at 300 equally-spaced points in the interval [0, 30] (panel (Figure 1a, density curves).

To compute the inequality in the expected infectiousness, we followed the approach from [12, SI, Sec. 2.2.5]. More specifically, we used the relation 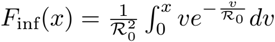 which is the expected fraction of all infections due to those who caused up to *x* infections. The expected fraction of all infections due to those who caused more than *x* infections would then be 1 − *F*_inf_(*x*), which is equal to 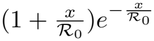 . The expected population fraction of the individuals who caused more than *x* infections is 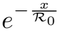 (Figure 1b, expected infectiousness). We used a similar approach to compute the inequality in the secondary case distribution: We used the relation 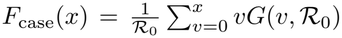 which is the expected fraction of all cases caused by those who generated up to, and including, *x* secondary cases. Here *G*(*v, R*_0_) is a geometric distribution with mean *R*_0_. The expected fraction of all cases due to those who caused more than *x* secondary cases equals 1 *− F*_case_(*x*). The population fraction of individuals causing more than *x* secondary cases would be 1 *− P* (*x*), where *P* () is the cumulative distribution function of a geometric distribution with mean *R*_0_ (Figure 1b, secondary cases).

### 4.4 Calculation of cohort mean and within-cohort variance of secondary cases

We used the rescaled SIR model (7) to compute the mean and variance of the expected secondary cases *C*(*·*). The SIR differential equations were numerically solved for the proportions of susceptible *x* and infectious *y* at a set of predefined equally-spaced time points.

We associated a (forward) cohort with each of the first 60% of time points. We used a 60% cutoff so that all included cohorts had almost fully recovered by the end of the simulation period, as verified numerically. We then calculated the cohort-specific mean and variance. More specifically, for the forward cohort infected at time point *τ*, the mean, *µ*(*τ*), and variance, *σ*^2^(*τ*), of the expected secondary are:

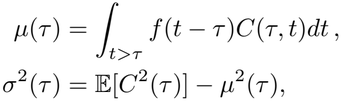

where

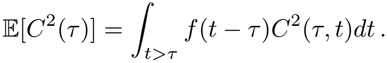

Here *f* () is the probability density function of the infectious period, and *C*(*τ, t*) is the expected number of secondary cases caused by the interval cohort *c*_*τ,t*_ (see Section 2.3 for definition). To calculate the quantity *C*(*τ, t*), the time series (*t, R* _0_*x*(*t*)) was linearly interpolated, and the result was numerically integrated from *τ* to *t*.

The cohort mean of expected secondary cases, *µ*(*τ*), was then computed by integrating of interval cohorts’ expected numbers of secondary cases weighted by the infectious period density function. Similarly, E[*C*^2^(*τ*)] was calculated for the forward cohort *τ* . We then computed the variance *σ*^2^(*τ*) by subtracting the square of the cohort-specific mean *µ*^2^(*τ*) from E[*C*^2^(*τ*)].Figure 4b was generated using this approach. The unweighted within-cohort dispersion for forward cohort *τ* was computed by dividing the variance *σ*^2^(*τ*) by *µ*^2^(*τ*) (Figure 4c).

To compute the mean expected secondary cases, *C*, over the entire epidemic, we separately applied linear interpolation to the incidence time series, (*τ, i*(*τ*) = *R R*_0_*x*(*τ*)*y*(*τ*)), and cohort mean expected secondary cases time series, (*τ, µ*(*τ*)). We then numerically integrated the product of these two interpolated time series and normalized the result by the final size, ∫ _*τ>* 0_ *i*(*τ*)*dτ* .

The within-cohort component of the variance was computed by numerically integrating the interpolated cohorts’ variance weighted by the interpolated incidence. The result was then normalized by the final size:

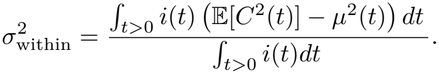

To compute the between-cohort component of variance, we numerically integrated the square of the interpolated cohort-specific mean, weighted by the interpolated incidence, and normalized it by the final size. Subtracting *C*^2^ then yielded the between-cohort component of variance.

The total variance was computed by summing the between- and within-cohort components of variance.

All numerical integrations were done using lsoda method with the R package deSolve. The linear interpolations were conducted using the R function approxfun. All simulations were carried out using R 4.5.0. Code for all numerical simulations can be found on Github: https://github.com/dushoff/emStaging. We solved all integrals across a range of values for *R*_0_, using the starting values *y*_0_ = 10^*−*9^; *x*_0_ = 1 *−* 10^*−*9^ to represent the limiting case in which there are no exogenous cases.

## Data Availability

All code and simulation files can be found at https://github.com/dushoff/emStaging

## Data and code availability

All code and simulation files can be found at https://github.com/dushoff/emStaging.

### Acknowledgments

S.J.B. and J.S.W. are investigators at the University of Maryland-Institute for Health Computing, which is supported by funding from Montgomery County, Maryland and The University of Maryland Strategic Partnership: MPowering the State, a formal collaboration between the University of Maryland, College Park and the University of Maryland, Baltimore. J.S.W acknowledges support from Simons Foundation grant MPS-SIP-00930382. This research was enabled in part by support provided by the Digital Research Alliance of Canada (alliancecan.ca).

## Supplementary Information for

### S.1 Analytical Results

The standard SIR model is described by the following equations:

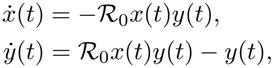

where *x*(*t*) and *y*(*t*) denote the proportions of susceptible and infectious individuals, respectively, and time is in units of mean infectious period. Let *i*(*t*) denote the incidence *R*_0_*x*(*t*)*y*(*t*), and define the final size of the epidemic as follows:

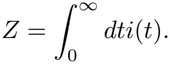

The probability density function of the infectious period is denoted by *f* (·), and the corresponding survival function is

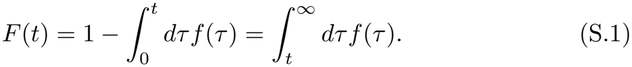

Note that *F* (*t*) is the probability that an individual infected at time 0 remains infectious after time *t*.

As detailed in Section 2.3, the expected number of secondary cases caused by the interval cohort *c*_*τ,ρ*_ that became infectious at time *τ* and recovered at time *ρ* is

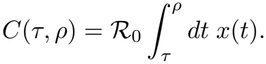

The *k*^th^ weighted raw moment of *C* is then

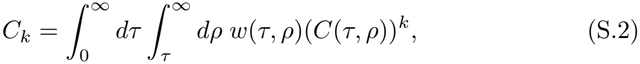

where *w*(*τ, ρ*) = *i*(*τ*)*f* (*ρ − τ*) is the size of the interval cohort *c*_*τ,ρ*_ .

In Section S.1.1, we prove the following statement: The excess variation of the individual case reproductive number in the standard SIR model outbreak is 1, provided that the initial infectious cohort is infinitesimal. Mathematically, this amounts to proving that

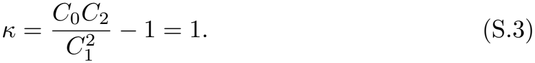

#### S.1.1 Proof

We set out to calculate *C*_0_, *C*_1_ and *C*_2_. We can calculate *C*_0_ and *C*_1_ without making any assumptions about the functional form of *f* (we do not have to assume that the infectious period exponentially distributed). To obtain *C*_2_, we assume that the distribution is memoryless, i.e., *F* (*a* + *b*) = *F* (*a*)*F* (*b*). Calculating each of these moments involves multiple manipulations of the limits of the integrals. To proceed, we assume that size of the initial infectious cohort is negligible.

From Equation S.2, we have

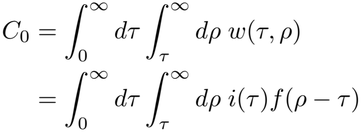

Substituting *ω* + *τ* for *ρ* in the inner integral, results in

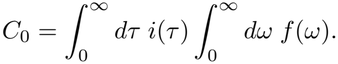

Since *f* a distribution with a nonnegative support, the inner integral is equal to 1. It follows that

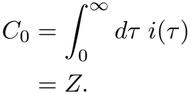

The next step is to prove that *C*_1_ = *Z*. From Equation S.2, we have

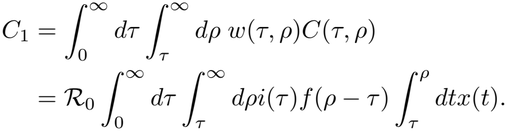

Instead of first integrating over *t* from *τ* to *ρ* and then over *ρ* from *τ* to, we can first integrate over *ρ* from *t* to and then integrate over *t* from *τ* to, i.e.,

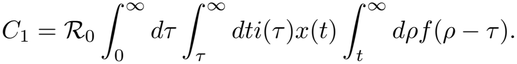

Substitute *ρ* = *ω* + *τ* in the innermost integral and using Equation S.1, we get

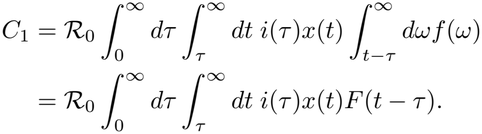

By interchanging the order of the integrals once more, we have

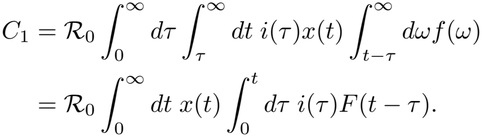

Note that the inner integrand counts the number of individuals that entered the infectious state at some positive time *τ < t* and remain infectious at time *t*. Assuming an infinitesimal initial infectious cohort, when we integrate over all times *τ < t*, we get the number of individuals who are infections at time *t*, i.e., *y*(*t*). So,

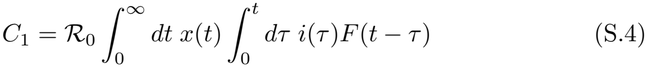

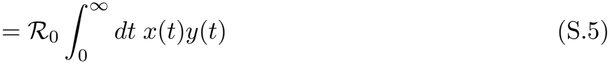

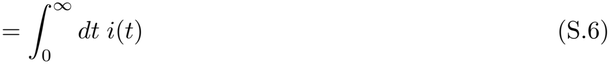

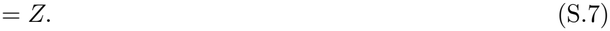

The next step is to show *C*_2_ = 2*Z*. Given Equation S.2, we have

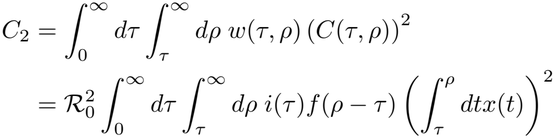

Now we break down the square integral into a double integral

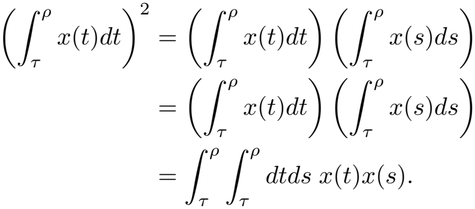

In *t − s* space, this is integrating *x*(*t*)*x*(*s*) over a square region whose vertices are (*τ, τ*), (*τ, ρ*), (*ρ, τ*) and (*ρ, ρ*). This is twice the integral over the triangle

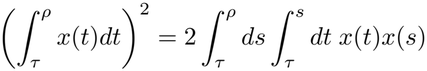

whose vertices are (*τ, τ*), (*τ, ρ*) and (*ρ, τ*). So,

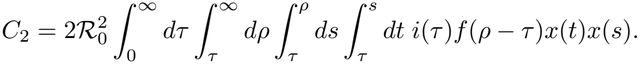

Plugging this back into the expression for *C*_2_ yields

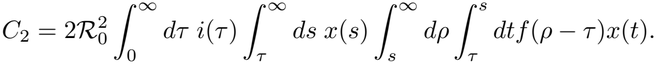

First, we reorder the integrals over *ρ* and *s* and change the limits appropriately

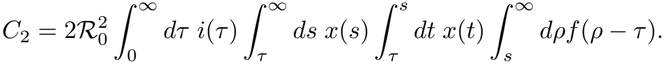

Next, we reorder the integrals over *ρ* and *t* to get

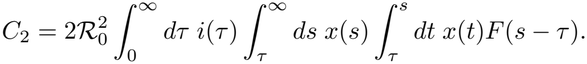

By substituting *ρ* = *ω* + *τ* and using Equation S.1, we get

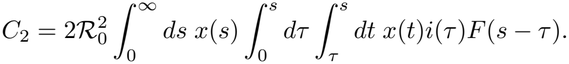

We then reorder the integrals over *τ* and *s*

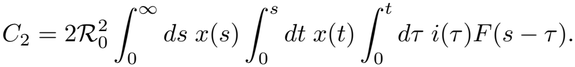

Swapping the integrals over *τ* and *t*, results in

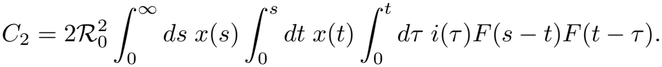

Now, we use the fact that the survival function *F* of an exponential distribution satisfies *F* (*a* + *b*) = *F* (*a*)*F* (*b*), and replace *F* (*s − τ*) with *F* (*s − t*)*F* (*t − τ*)

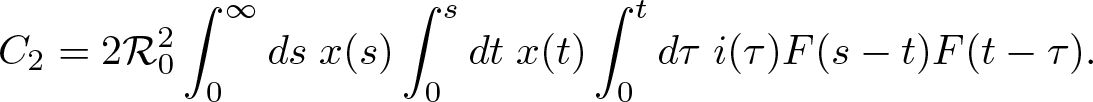

Next, we use the same argument we used to go from Equation S.4 to Equation S.5, which results in

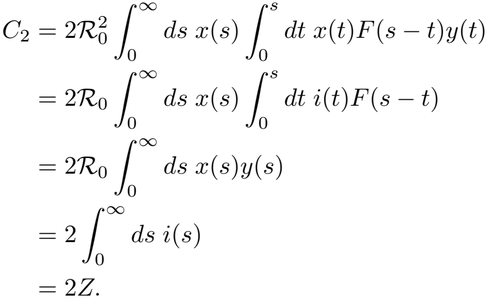

Finally, replacing *C*_0_, *C*_1_, and *C*_2_ with *Z, Z*, and 2*Z*, respectively, in Equation S.3 yields *κ* = 1.

#### S.1.2 Incorporating the Size of Initial Infections

Equation S.3 was shown to hold under the assumption of an infinitesimal initial infectious cohort. Here, we revisit the results while relaxing this assumption.

To account for the initial infections, we assume that the epidemic starts abruptly at time *t* = 0, i.e.,

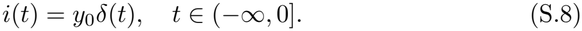

Here, *y*_0_ is the proportion of the population that were infectious at the beginning of the outbreak, *t* = 0, and *δ*(*t*) denotes the Dirac delta distribution [2]. Then, by accounting for the initial infectious cohort, the proportion of the infected population that is infectious at time *t* is

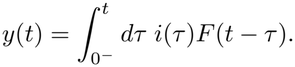

Here, the lower limit of the integral is 0^*−*^ to include the Dirac delta function at *t* = 0. The total weight *C*_0_ will now includes the initial infectious cohort size as well, i.e,

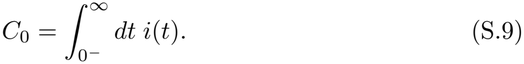

The first weighted raw moment *C*_1_ is given by

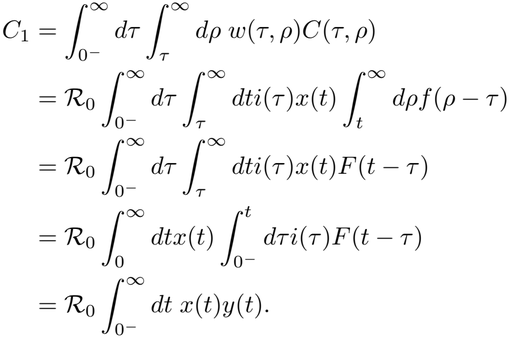

Given both *x*(*t*) and *y*(*t*) are bounded, without loss of generality, the lower limit can be replaced with 0^+^. Thus, the first weighted raw moment *C*_1_ becomes

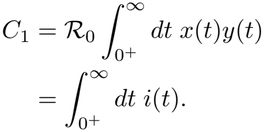

We note that *C*_1_ does not include the size of initial infectious cohort. Now we move on to recalculate *C*_2_, while taking the initial infectious cohort into account.

We have

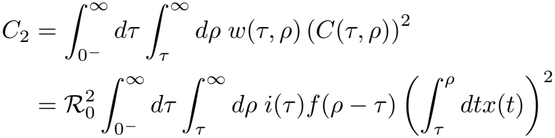

Using the same mathematical manipulation as in Section S.1.1, we obtain

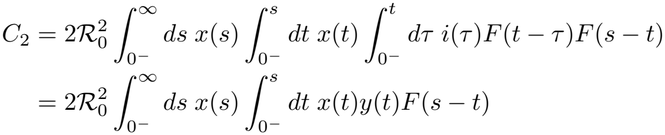

We split the innermost integral into two integrals

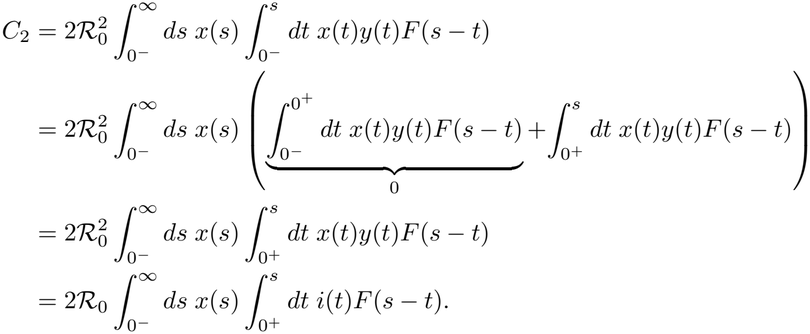

The innermost integral gives the proportion of the population who entered the infectious state at some time *t >* 0 and remain infectious at time *s*. Hence, it equals the total proportion infectious at time *s* minus the proportion of the population who were infectious at *t* = 0 and remain infectious at time *s*, i.e., *y*(*s*) *− y*_0_*F* (*s*). Having replaced the innermost integral with *y*(*s*) *− y*_0_*F* (*s*), we obtain

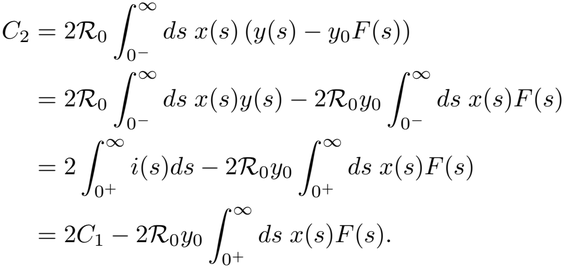

Note that changing the lower limit of integration from 0^*−*^ to 0^+^ does not affect the result because the integrand is bounded. The second term in the right hand side is the twice the expected secondary cases of the initial cohort. Here we notice that the second weighted raw moment is *dependent* on the key parameter *R*_0_. However, as the initial cohort size *y*_0_ tends to zero, (i) the difference between *C*_2_ and twice the first weighted raw moment 2*C*_1_ approaches zero; (ii) the total weight *C*_0_ (S.9) converges to *C*_1_. Thus, Equation S.3 is retrieved.

